# Hospital Trainees’ Worries, Perceived Sufficiency of Information and Reported Psychological Health During The COVID-19 Pandemic

**DOI:** 10.1101/2020.07.22.20158311

**Authors:** Nikoo Aziminia, Aria Khani, Colette Smith, Ameet Bakhai, Clifford Lisk

## Abstract

**Introduction:** The COVID-19 pandemic has been unsurpassed in clinical severity or infectivity since the 1918 Spanish influenza pandemic and continues to impact the world. During the A/H1N1 influenza pandemic, healthcare workers presented frequent concerns regarding their ownand their families’ health, as well as high levels of psychological distress.

**Objectives:** To assess hospital trainees ‘concerns, perceived sufficiency of information, behaviour and reported psychological health during the COVID-19 pandemic in the NHS

**Design:** Cross-sectional 39-point questionnaire study conducted in May 2020

**Setting:** A large NHS foundation trust in London

**Participants:** 204 hospital trainee doctors

**Outcome measures:** Quantitative analysis of trainees’ worries and concerns while working during the COVID-19 pandemic were assessed across 8 domains: trainee demographics; concerns and worries regarding COVID-19; perceived sufficiency of information about the COVID-19 pandemic; social distancing; use of personal protective equipment (PPE) and training in PPE; COVID-19 acquisition and risk; reported psychological health; and medical education.

**Results:** 91.7% looked after COVID-19 patients. 91.6% were worried about COVID-19; the most frequent concern was that of family and friends dying from COVID-19 (74.6%). 22.2% reported being infected with COVID-19. 6.8% of trainees were so concerned about COVID-19 infection that they would avoid going to work. Perceived sufficiency of information about COVID-19 was moderately high. 25.9% reported that they were able to socially distance at work compared to 94.4% able to socially distance outside work. 98.2% reported using PPE and 24.7% were confident the provided PPE protected them against infection with COVID-19. 41.9% reported that their psychological health had been affected by their work with the commonest being anxiety (56.6%), emotional distress (50.9%) and burnout (37.7%). 95.6% felt it is important to have a service that provides psychological support during this pandemic and 62.5% reported they would consider using this at work.

**Conclusions:** A significant proportion of hospital trainees are worried about the COVID-19 pandemic with high levels of reported psychological distress. Given that almost a third would not use psychological support services at work, hospital leaders and liaison psychiatry need to explore the reasons for not wanting to use services at work and highlight the provision of psychological services provided outside work such that provided by the London deaneries professional support unit (PSU). Seeking solutions to support trainee wellbeing in addition to this, such as larger offices, adequate rest facilities and alternative methods of teaching, with their input would enable empowerment of trainees and improve their health and morale while working in a pandemic.

## Introduction

In December 2019, the first reports of a cluster of cases of pneumonia of an unknown aetiology emerged from Wuhan, China.(1) As other respiratory pathogens such as SARS-CoV, MERS-CoV and influenza were excluded as causes, a novel coronavirus was subsequently identified and named SARS CoV-2, the disease caused by this being named COVID-19. The day after the genetic sequence of COVID-19 was publicly shared by China in January 2020, the first case of COVID-19 outside of China was identified in Thailand.(2) By March 2020, the WHO had declared a pandemic.(3)

As of 22^nd^ July 2020, there have been 14,731,563 confirmed cases of COVID-19 and 611,284 deaths reported to WHO globally.In the UK to date there have been 295,376 confirmed cases of COVID-19, the ninth highest case burden in the world and the highest in Europe, with45,312 deaths.(4) Of the UK deaths, 181 were NHS workers and 131 were social care workers, including several doctors.(5,6)To date, it has been unsurpassed in terms of clinical severity and transmissibility since the 1918 Spanish influenza pandemic.(7)

A crucial element of the UK Department of Health’s pandemic preparation following the H1N1 pandemic in 2009 wasa containment strategy.(8) While it is arguable how effective such a strategy would be at a point where the most populous country of the world had already succumbed to an epidemic and infection has spread to every continent, preventing healthcare systems from being overwhelmed amid increased demands has been a priority.(9) Protecting the NHS was central to the UK Government’s message to the public at the onset of national lockdown(10). Inevitably, hospital practices changed significantly during the COVID-19 pandemic in order to effectively manage patients infected with SARS CoV-2 and to limit transmission.

The NHS is the 5^th^ biggest employer in the world, comprising of over 1 million full-time staff of whom 122,031 are doctors(11,12). Hospital trainees, doctors in postgraduate training working in the hospital setting, have been on the frontline of the COVID-19 pandemic. During the previous pandemic of this scale, H1N1, hospital workers reported significant concerns regarding their own health and that of their families, with potential impact on their ability to perform their duties(13). The aim of our study was to assess the concerns of hospital trainees and the effects, perceived or otherwise, of working during this global health COVID-19 crisis on their training, their ways of working, and their physical and psychological wellbeing.

## Methodology

This cross-sectional questionnaire study was carried out between the 1^st^ and 31^st^ of May 2020 at Barnet Hospital and the Royal Free hospital (part of the Royal Free London group of hospitals within the foundation trust) United Kingdom. Barnet hospital (459 beds) and The Royal Free hospital (839 beds) are a district general hospital and a teaching hospital providing secondary and tertiary care for a population of over 900,000 people. At the time of writing this paper, 5859 patients with COVID-19 have been admitted at both hospitals with 639 deaths.

A 39-item questionnaire was developed by the authors, adapted from a previous questionnaire by Goulia et al. with permission, to assess hospital doctors’ in training anxieties, worries and concerns during the pandemic; their perceived sufficiency of information concerning COVID-19; their intended behaviour; use of PPE; reported psychological health; experience of medical education; and their experience of self-isolation.(13)Two items were scored on a 9-point Likert scale from strongly agree (9) to strongly disagree (1) and one was scored on a 9-point Likert scale from extremely worried (1) to extremely unworried (9) (Figures 2 and 8). The remaining items were dichotomous.

**Figure 1:**
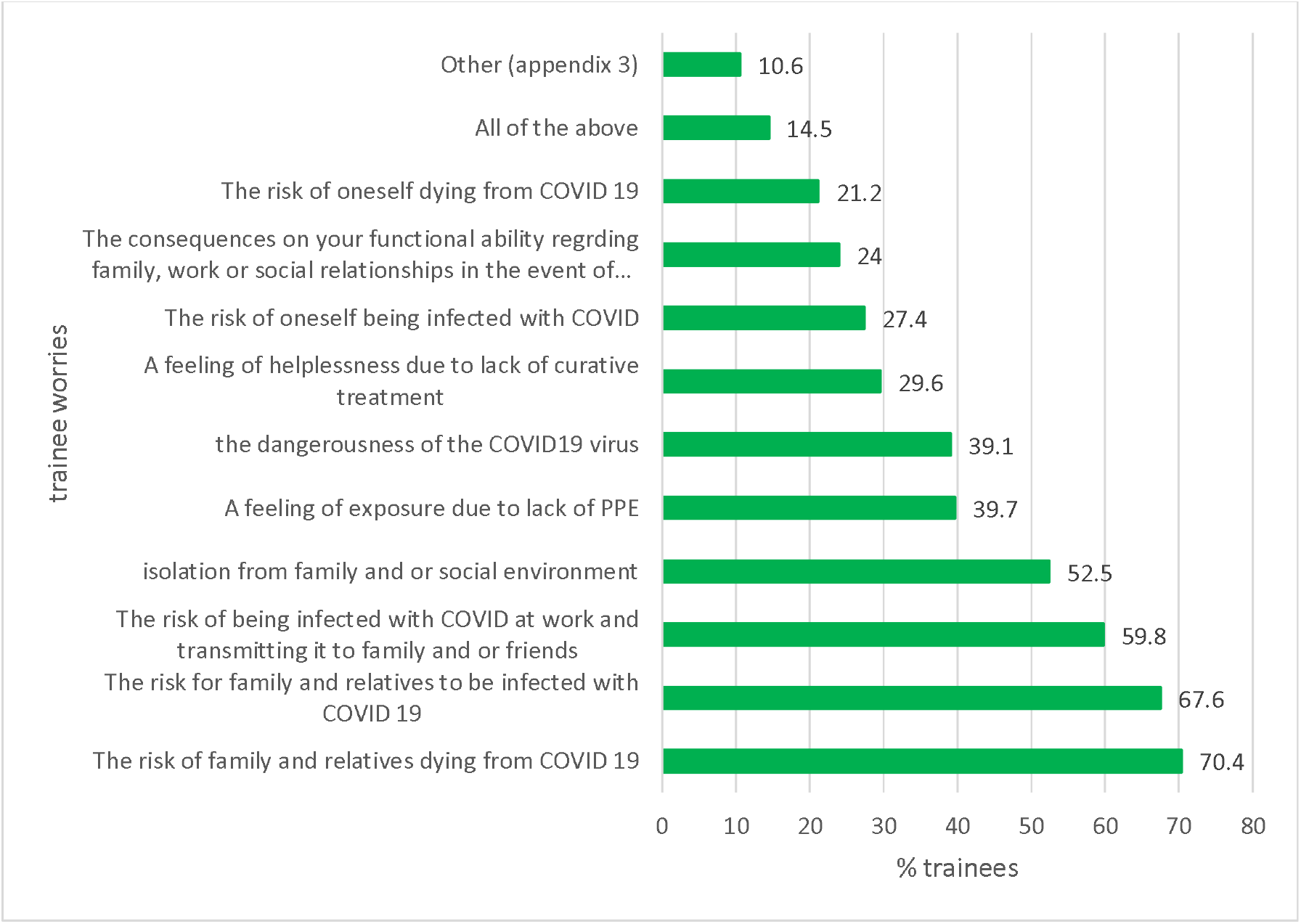
Trainee worries during the COVID-19 pandemic.

**Figure 2:**
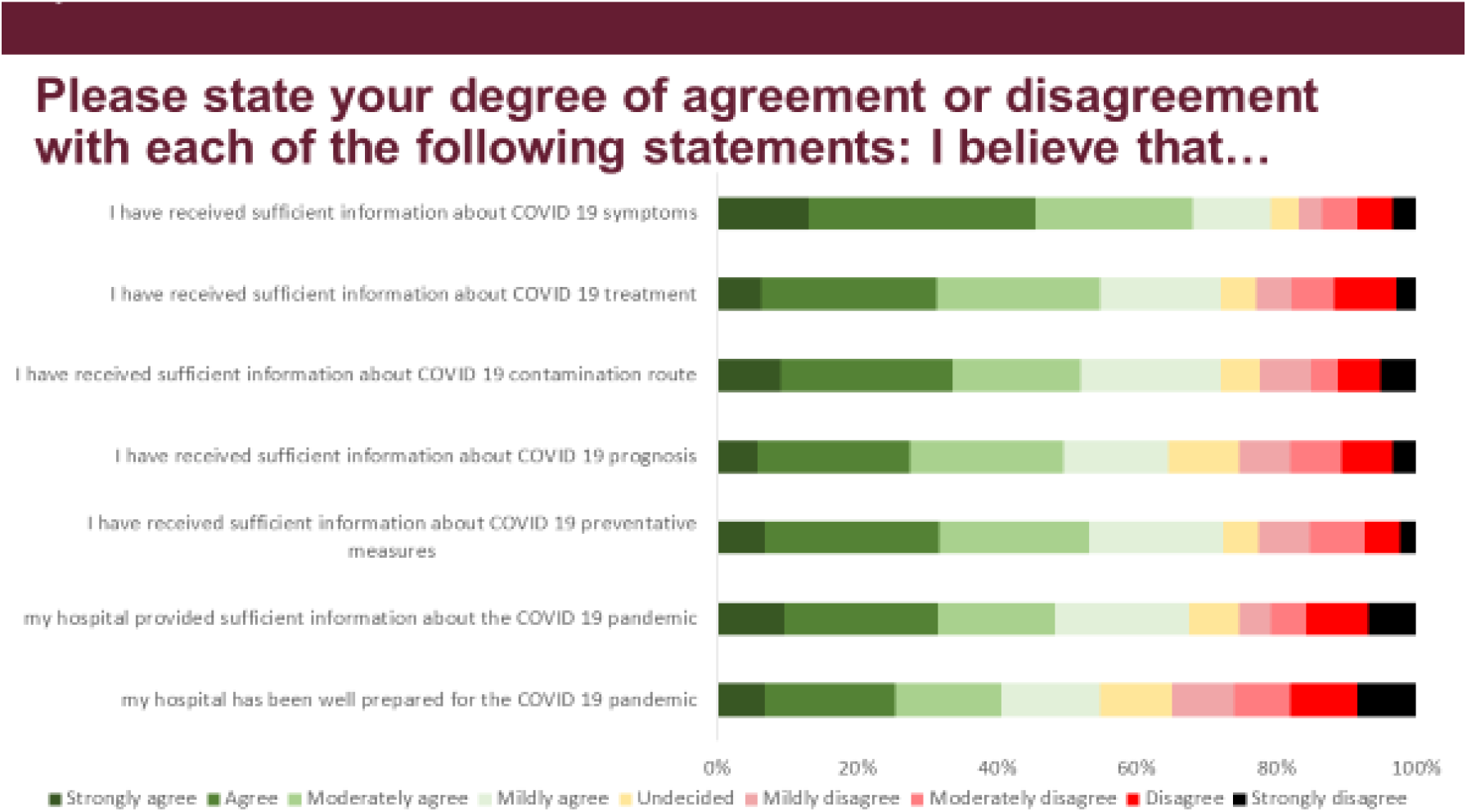
Perceived sufficiency of information about the COVID-19 pandemic and about hospital preparedness.

Items were grouped into 8 domains; demographics and professional information; concerns regarding COVID-19; perceived sufficiency of information about COVID-19; social distancing; use of personal protective equipment; COVID-19 acquisition and risk; mental health and medical education. Denominators vary depending on how many trainees answered each question and the lowest number of responses to any question was 159.

### 1. Recruitment

Hospital doctors in training in all hospital departments and clinical units were asked to participate in this study via a Survey Monkey questionnaire (Appendix 1a). This was distributed by email via the administrative staff at the Postgraduate Medical Education Centre at Barnet Hospital. The email explained the purpose of the study and its aims. The first page of the questionnaire comprised further information and informed consent. All the procedures followed were in keeping with ethical standards (world medical association Helsinki declaration). The UK Health Research Authority indicated that the project did not require ethical review by an NHS/HSC or social care research ethics committee or management committee through the NHS/HSC research and development office. The project was discussed with the Royal Free Hospital R&D office who stated that no further approvals were required. Reminder emails were sent twice, one week apart to all hospital doctors in training at Barnet Hospital and Royal Free Hospital by the administrative team at the Barnet Hospital Postgraduate Medical Education Centre.

### 2. Statistical Analysis

All statistical analyseswere performed using the Stata version 14. Summary statistics for all variables were calculated using a complete case analysis. Questionnaire responses were stratified according to: (i) whether the trainee doctor reported or did not report any personal mental health concerns (ii) whether they were or were not redeployed during the pandemic and (iii) whether they had or had not been able to socially distance at both home and work. Chi-squared analysis, Fishers Exact and Mann-Whitney tests were performed to assess the strengths of associations.

## Results

### 1. Demographics and professional information

Of 485 hospital doctors in training sent the survey by email, 204 completed the questionnaire (42.1%response rate). There were 123 women (60.3%), with the most common ethnicity being Caucasian (49.5%, 101) followed by Asian (24.5%, 50) (Table 1). The majority (91.2%; 186) had been looking after COVID-19 patients and 46.1% (94) were redeployed during the pandemic. Trainees were redeployed to the acute COVID-19 rota from a wide variety of areas including academic placements, dermatology, ENT, final year medical students, general practice, general surgery, histopathology, maxillofacial surgery, medical education, psychiatry, urology and virology (Appendix 1).

**Table 1:** Demographics and professional characteristics.

|  |  |
| --- | --- |
| **Where do you work? \*** | **N (%)** |
| Barnet Hospital | 104 (51.0%) |
| Royal Free Hospital | 84 (41.2%) |
| Chase Farm | 4 (2.0%) |
| GP based | 7 (3.4%) |
| Other | 9 (4.4%) |
| **What is your gender?** |  |
| Female | 123 (60.3%) |
| Male | 80 (39.2%) |
| Other/prefer not to say | 1 (0.5%) |
| **What is your ethnicity?** |  |
| Asian/Asian British | 50 (24.5%) |
| Black/Black British | 6 (2.9%) |
| Chinese/Chinese British | 9 (4.4%) |
| Middle Eastern/Middle Eastern British | 10 (4.9%) |
| Mixed race – other | 14 (6.9%) |
| **Other ethnic group** | 5 (2.5%) |
| White – British, Irish, Other | 101 (49.5%) |
| Prefer not to say | 9 (4.4%) |
| **What level of practice are you currently at?** |  |
| Core medical training Y1 | 8 (3.9%) |
| Foundation Y1 | 32 (15.7%) |
| Foundation Y2 | 30 (14.7%) |
| Internal medicine Y1 | 9 (4.4%) |
| Specialist Registrar | 76 (37.3%) |
| Other (GP trainees, Surgical trainees, Emergency Medicine Trainees, clinical fellows and Foundation Year 3 doctors) | 49 (24.0%) |
| **Have you been involved in looking after COVID-19 patients?** |  |
| Yes | 186 (91.2%) |
| No | 18 (8.8%) |
| **Have you been re-deployed during this COVID-19 pandemic?** |  |
| Yes | 94 (46.1%) |
| No | 110 (53.9%) |

\*4 people listed two places of work

### 2. Concerns and worries regarding COVID 19

In total, 91.6% (164/179) of trainees were worried about COVID-19 (Figure 1). The three topmost worries for trainees were the risk of families and friends dying from COVID-19, the risk of family and friends being infected with COVID-19 and the risk of being infected at work and transmitting it to family and or friends (Table 5). Trainees were less worried about themselves being infected with COVID-19, dying from COVID-19, or the consequences on their functional ability regarding family, work or social relationships in the event of being infected with COVID-19. Concerns about isolation from family and or the social environment and about a feeling of exposure due to lack of PPE were moderately high (Figure 1). Worries were higher among those reporting mental health concerns (97.7% vs 83.8%, p= 0.002), but similar for those who were redeployed (95.2% vs 89.4%, p=0.15) and those who were socially distanced (95.1% vs 90.8%, p=0.38). Other trainee worries were expressed in free text responses(Appendix 2).

### 3. Perceived sufficiency of information about the COVID-19 pandemic

The degree of trainees’ perceived sufficiency of information regarding COVID-19 symptoms, prognosis, contagion route, treatment, preventative measures, provision of information relating to the COVID-19 pandemic by the hospital, and hospital preparedness was high, with agreement ranging from 50-80% (Figure 2). 88% of trainees agreed they had received enough information about COVID-19 symptoms. 54.8%(158) of trainees agreed their hospital was well prepared for the pandemic. Perceived sufficiency of information was broadly similar between trainees with no reported mental health issues and those with reported mental health issues, those redeployed and those not redeployed and those who socially distanced and those who didn’t socially distance (Tables 9-11). There was significance variation between redeployed and non-redeployed trainees in agreement with regards to the statements “I have received sufficient information about COVID-19 treatment” (Median IQR 4 vs 3, p=0.0006) and “I have received sufficient information about COVID- 19 preventative measures” (Median IQR 4bs 3, p=0.001).

## 4. Social distancing

25.9% (42) of trainees implemented the recommended social distancing at work whilst the majority (94.4%, 152) implemented this outside work (Figure 3). Of those who had been redeployed, 50% did not socially distance at work compared to 33.3% who did not (p=0.062). There were similar rates of worry about COVID-19 amongst trainees who did not socially distance and those who did (90.8% vs 98.1%; p=0.38). 100% of trainees practising social distancing used personal protective equipment (PPE) compared with 92.9% of those who did (p=0.003).

**Figure 3:**
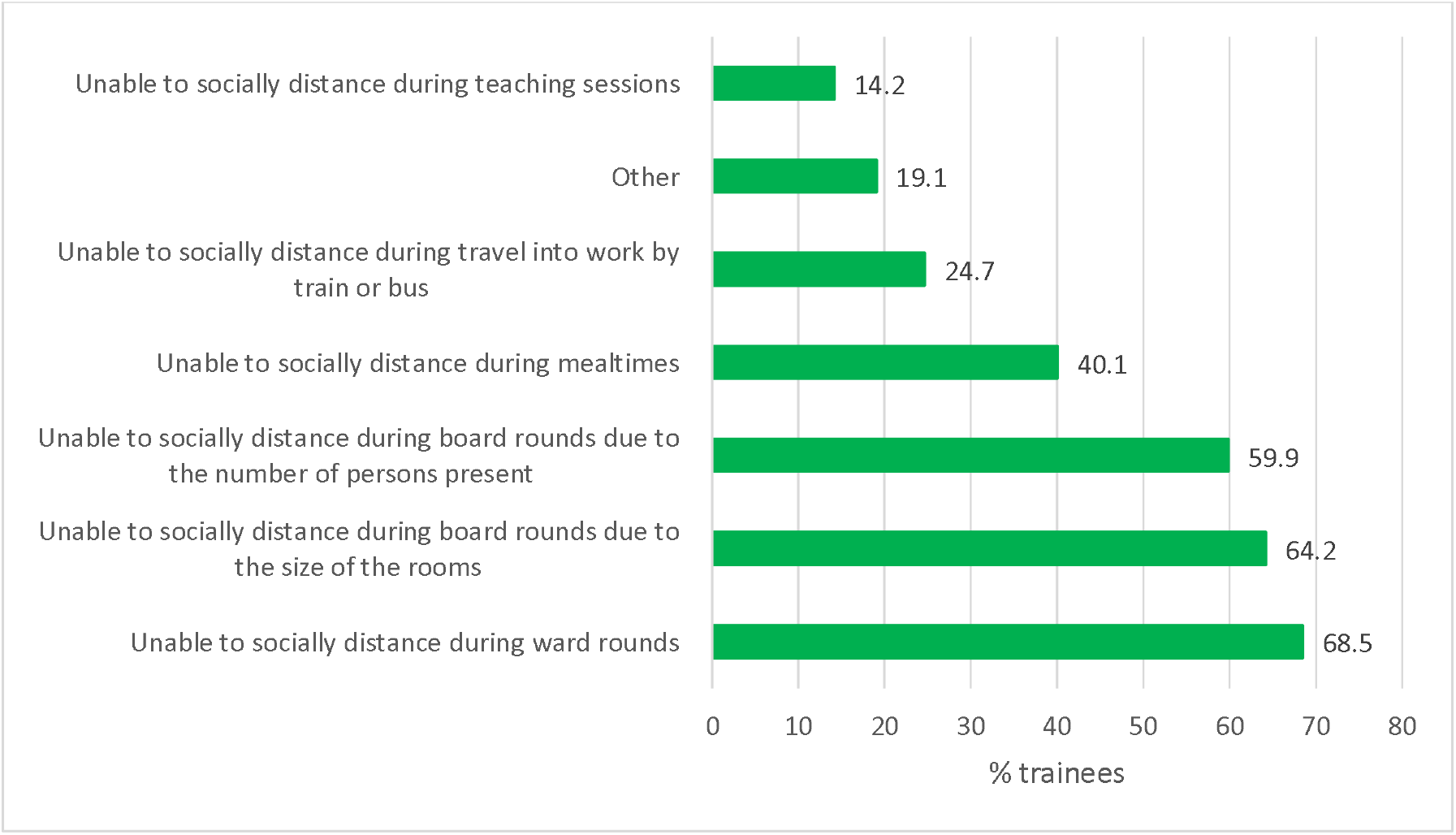
Social distancing at work and outside work.

The commonest reasons cited for not being able to socially distance at work were ward rounds (68.5%, 105) and board rounds due to the size of the rooms (64.2%,99) and due to the number of persons present (59.9%, 92). (Figure 4). Other comments highlighting the logistical difficulties of maintaining social distancing at work were raised in free text responses (Appendix 3). The commonest suggested trainee solutions to enable social distancing at work were larger offices (82.1%,129), more computers (74.1%,116) and larger recreational spaces (60.5%,95). Remote ward rounds and remote teaching were indicated as solutions by 38.2% (60) and 35.7% (56) of trainees respectively.

**Figure 4:**
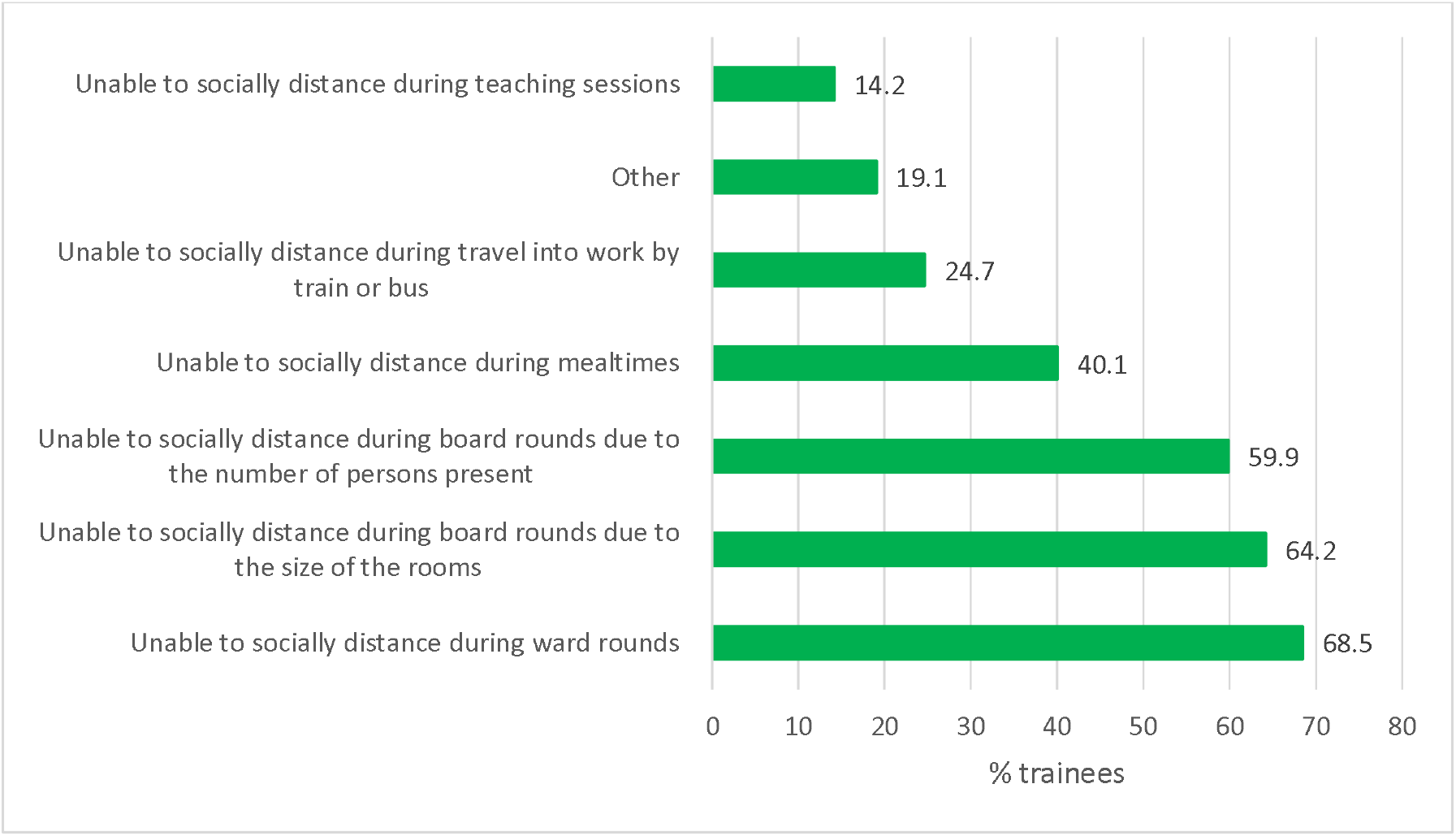
Reasons why trainees cannot socially distance at work (n=162)

## 5. Use of personal protective equipment (PPE) and training in PPE

98.2% (159/163) of trainees used PPE whilst caring for patients with COVID-19 and this was comparable for those with and with mental health concerns (96.6% vs 100% p=0.13), those not redeployed and those redeployed (97.7% vs 98.7%, p=0.67),those who socially distanced and those who did not socially distance (100% vs 92.9%, p=0.003). Of these, the most commonly used PPE were disposable gloves (93.8%, 152), disposable plastic aprons (91.4%,148) and scrubs (87.7%, 142). 68.9%, (111) used FFP3 masks, and46.6% (75) used fluid resistant type IIR surgical face masks and 59% (95) used non-fluid resistant surgical masks.64% (103) reported using face shields(Figure 6).

**Figure 6:**
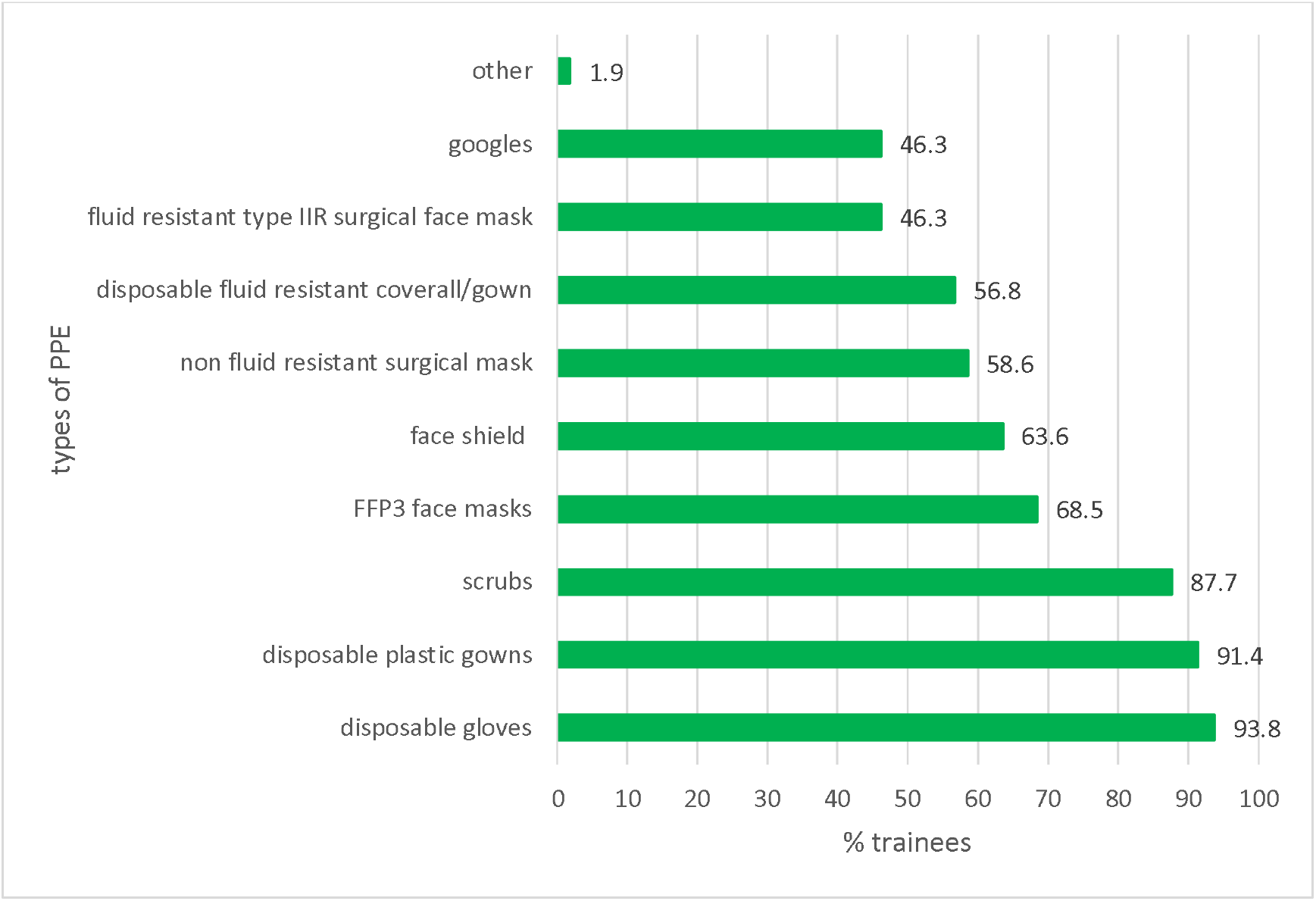
PPE used by trainees.

70.8% (114) of trainees had training in donning and doffing. This was comparable for trainees without and with mental health concerns (76.2 vs 68.2; p=0.25), those not redeployed and those redeployed (66.7% vs 75.7%, p=0.21) and those not socially distancing and those social distancing (70.8% vs 70.7%, p=0.99). Of those who have had this training, 56.6% (89) agreed this training was sufficient. 24.7% (39) were confident that the PPE they were wearing at work protected them from being infected with COVID-19. 66% (107) were confident that they were following the recommended COVID-19 infection control guidelines at work and 61.7% (100) were confident they were donning and doffing the recommended PPE correctly at work (Figure 7).

**Figure 7:**
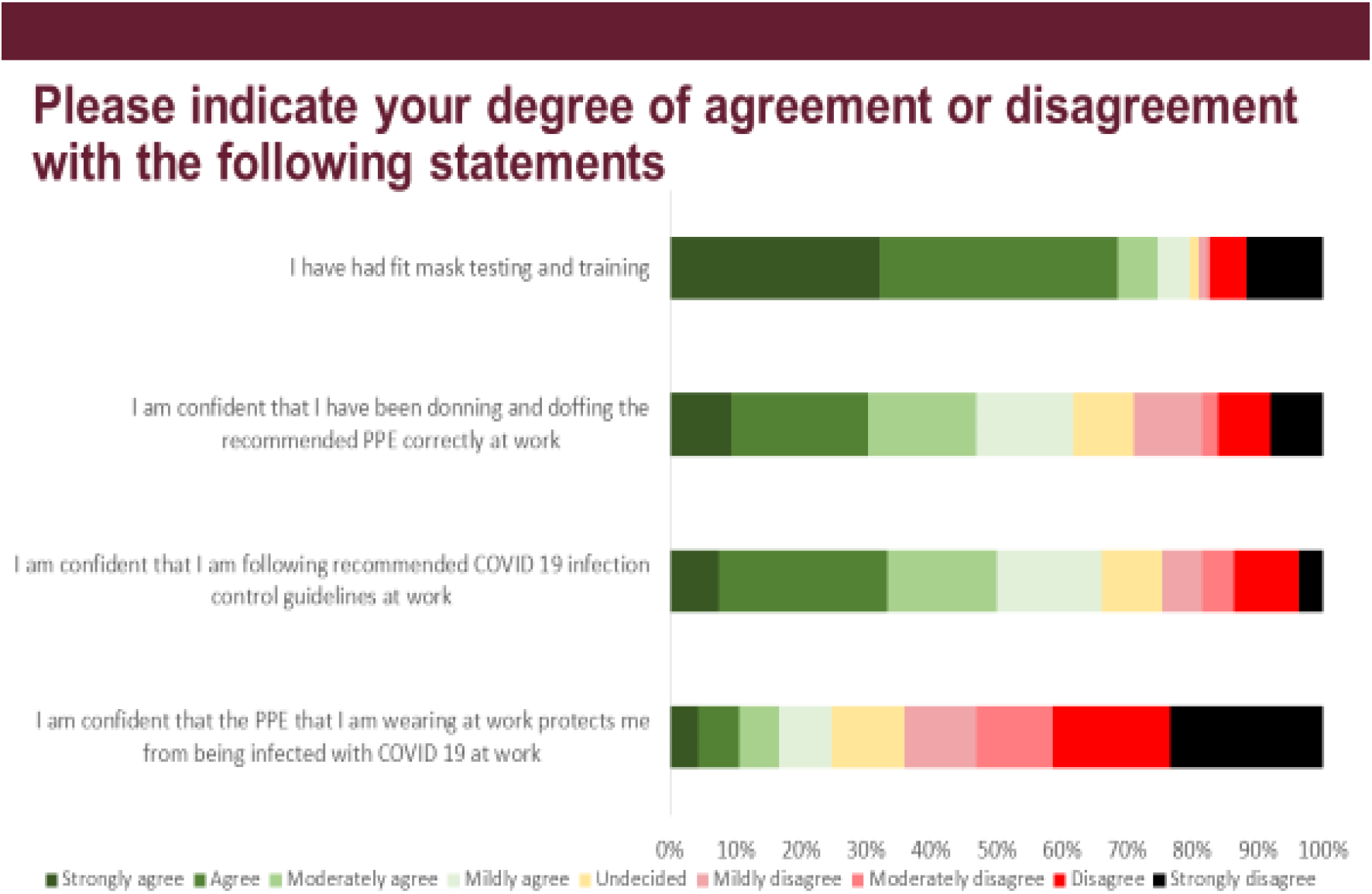
Trainees views with regards to PPE.

## 6. COVID-19 acquisition and risk

22.2%(36) of trainees reporting having been infected with COVID-19. 25% (17) and 20.5% (18) of trainees with no mental health concerns and mental health concerns had been infected with COVID- 19 respectively (p=0.063). 28.8% (21) of redeployed and 17.1% (15) of non-redeployed trainees had been infected with COVID-19 respectively (p=0.15), whereas reported infection rates were comparable for trainees who had not socially distanced and those who had (23.5%(28) vs 19.1% (8), p=0.64). Of those trainees who were unsure or reported they had not been infected (n=132), 84.1%(110) perceived their risk of being infected with COVID-19 as high. The perceived risk of being infected with COVID-19 was high across all the groups; those with and without mental health concerns 83.8% (62) and 85.2% (46) respectively); those redeployed and not redeployed (85.3%, 52 and 83.1%,59 respectively); and those who did and did not socially distance (88.8%,32 and 82.2%, 79 respectively)(Figure 8). 6.8% (11) of trainees were so concerned about contracting COVID-19 that they would avoid going into work. Trainees with no mental health concerns (92.5%,62) were more likely to avoid going into work than those with mental health concerns (71.9%,64).

**Figure 8:**
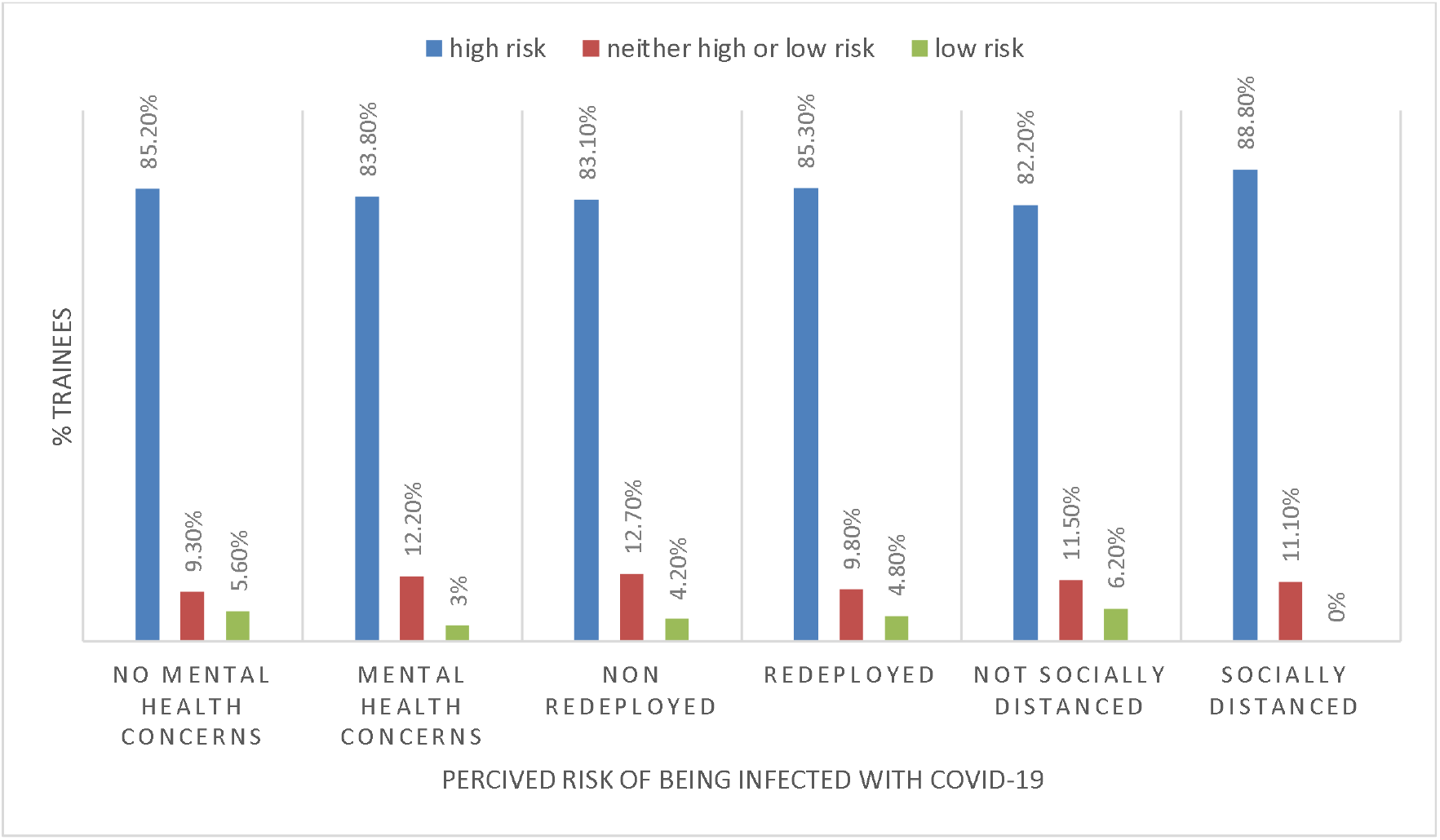
Trainees’perceived risk of being infected.

## 7. Reported psychological health

41.9% (67/160) of trainees reported concerns about their psychological health. The commonest reported concerns were anxiety (37.5%,25), emotional distress (33.8%,23) and burnout (25%,17) (Figure 9). 56.9% (91)trainees felt anxious about a colleague falling ill at work. 95.6% (153) felt it was important to have psychological support services during the COVID-19 pandemic with62.5% (100) stating they would consider using those services. 77.5% (124) of trainees were aware of the wellbeing support currently available at work;the most commonly stated avenues of support being educational and clinical supervisors (83.1%,133 and 76.9%, 123 respectively).

**Figure 9:**
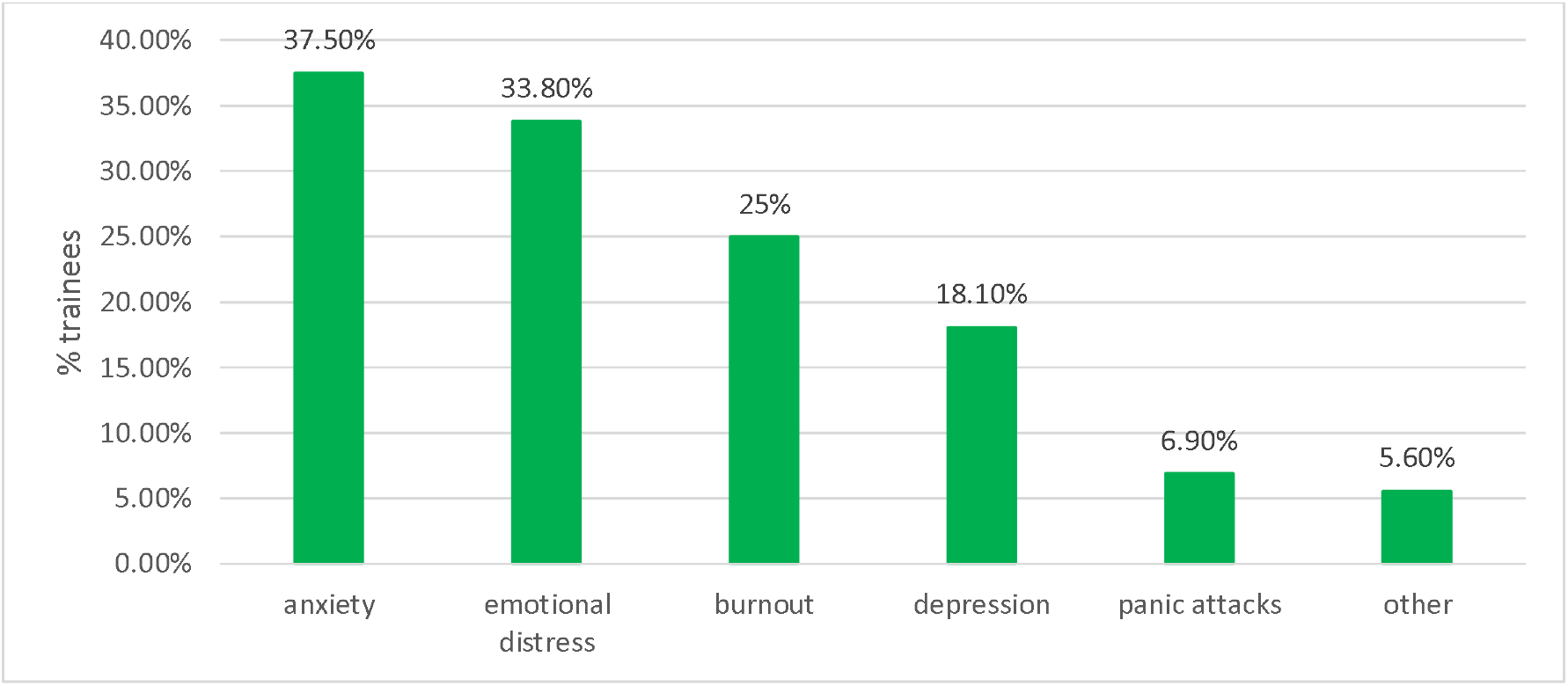
How trainees’ psychological health had been affected (n=160)

## 8. Medical Education

73% (116/159) of trainees were aware the delivery of medical education had changed during the COVID-19 pandemic. The most commonly reported alternative method of delivery of medical education was Microsoft teams (Figure 10). 61% (96) of trainees reported their commonest source of information regarding COVID-19as their employer. 53.5% (85) of trainees received information regarding COVID-19 via television news programmes and 50.9% (81) via daily government briefings.

**Figure 10:**
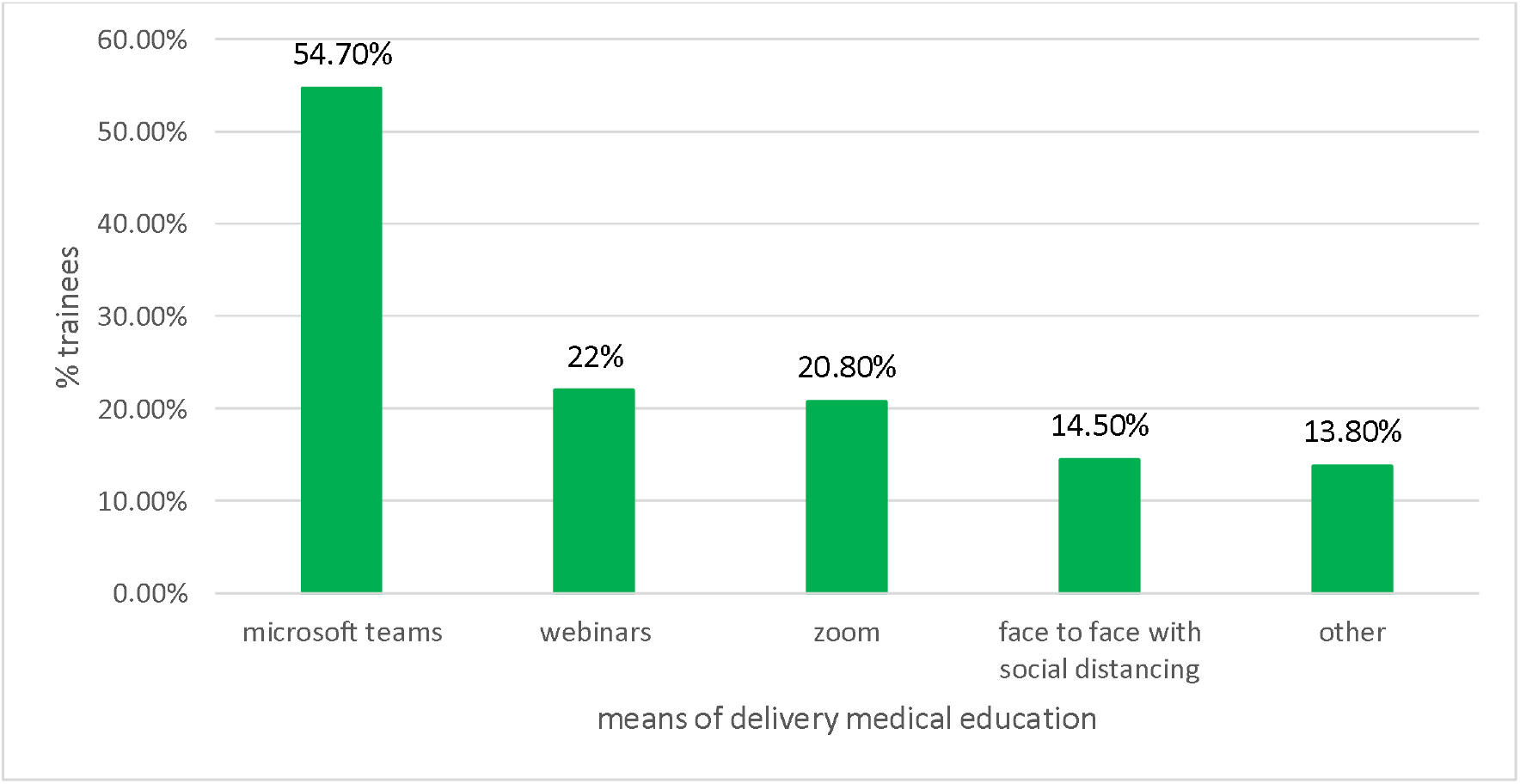
New methods of training (n=132)

## Discussion

Our study demonstrates that in May 2020 during the COVID-19 pandemic outbreak in the UK, in our hospitalmost trainees were concerned about the pandemic, with most reporting being “moderately” worried. When exploring potential reasons for this, the most recurrent concerns were for the wellbeing of family and relatives, above concerns for their own wellbeing. Due to the reality of working during the pandemic, the concern for loved ones gave rise to further concerns, namely with regards to social isolation measures andthe availability and quality of PPE.

Social isolation measures were more strictly adhered to outside of work rather than at work. This discrepancy appears to be driven by numerous factors, mainly the clinical environment itself serving as an obstacle to effective social distancing.Trainees also found it difficult to socially distances during non-clinical activities such as breaks, although many felt doing so was futile; some respondentsstated they were unable to maintain the recommended 2m distance in high-risk COVID- 19 areas, while others questioned the evidence to support socially distancing in clinical environments. This presents a significant issue. During an infectious disease outbreak, effective social distancing is an essential public health measure that has demonstrated the potential to limit the spread of infection and the resultant impact on society, without the use of vaccines or antimicrobials(14).This intervention has been projected to reduce the cost in terms of human life and also of resources; by reducing the disease burden, the risk of overwhelming hospital and intensive care unit capacity would possibly be reduced and the quality of care for other medical problems would be maintained.(15)In the presence of COVID-19, a disease with transmissibility and clinical severity comparable with the 1918 influenza epidemic(7),the inability to socially distance while dispatching clinical duties is highly concerning.Solutions to this problem were explored with participants; larger spaces at work were recommended by many, namely larger doctors’ offices and recreational spaces, to overcome barriers to social distancing. Investment into IT resources was another recommendation, whereby over three quarters agreed more computers would allow the minimum distance to be maintained as well as provisions for remote ward rounds, teaching, and computer software access.

Another key measure in hospitals to protect staff and limit the spread of infection is personal protective equipment (PPE). 98.2% of respondents stated they utilised PPE in their clinical practice. Of those who denied using PPE, reasons given included limited patient exposure in their clinical environment. 70.8% had received training in donning and doffing and 79.63% had undergone fit testing for masks, however there were mixed feelings about the sufficiency of their training, with 56.6% stating this was sufficient. While most respondents felt their hospital was well-prepared for the pandemic, 64.2% stated their disagreement with the notion that the PPE provided at work protected them from being infected, of whom over a third strongly disagreed. A significant area of concern voiced by participants was the issue of PPE. One participant reported having purchased their own goggles, visor, and scrubs throughout the pandemic and another suggested their infection with SARS CoV-2 was directly attributable to shortcomings in available PPE. PPE guidance as per Public Health England (PHE) has changed on a number of occasions throughout the pandemic and more recently been endorsed by the World Health Organisation (WHO).(16,17) However, subtle discrepancies exist. PHE guidance recommends the use of fluid-repellent surgical masks in clinical areas where aerosol-generating procedures are not regularly performed, while the WHO guidance recommends medical facemasks with no clarification as to the type to be used. Current evidence suggests SARS CoV-2 is spread through respiratory droplets, aerosols, direct or fomite contact, and faeco-oral transmission. While respiratory droplets tend to fall rapidly, SARS CoV-2 bio-aerosols have been shown to persist in air for at least 3 hours.(18) Coughing and sneezing can generate both droplets and aerosols, the majority of which are within the <20 µm range, while surgical facemasks may not offer protection against particles < 100 µm in size.(19) One study evaluating the protection against influenza bio-aerosols offered by a range of surgical facemasks found live virus was detectable behind all masks tested(20). In addition, in a global comparison of PPE guidance, PHE guidance was unique in its recommendation of a plastic apron as body protection and on conditional use of eye protection, whereas all other guidance used in the comparison, including that of WHO, recommended a long-sleeved gown or equivalent along with eye protection in all scenarios.(21) While all respondents with direct patient contact reporting using PPE, and that provided by the Trust was in keeping with PHE guidance, it is clear from our results that the PPE trainees used was proportionate toprovisions made; 91.4% reported using plastic aprons, whereas only 56.8% reported using disposable fluid resistant gowns or coveralls, as is the WHO guidance and that of numerous countries. Interestingly, there was some heterogeneity amongst the masks used by respondents; 46.3% reported using fluid-resistant surgical masks while 58.6% reported using non-fluid resistant surgical masks. Given non-fluid resistant surgical masks were not recommended in the PPE guidance any public health body, this begs the question as to why such a discrepancy was reported and whether all surgical masks were fluid resistant. While the death of healthcare professionals during this pandemic cannot be solely attributed to potential inefficacy of PPE as per PHE guidance, this remains a controversial issue on a national level.

Healthcare workers dying from COVID-19 is an issue that has inspired significant public concern. One of the aims of this study was to explore the physical and psychological wellbeing of hospital trainees. Almost a quarter (22.22%) of participants confirmed they had been infected with COVID-19 and over half (50.62%) were unsure, likely due to a lack of testing being both available (which was introduced for hospital stafffrom30^th^ March 2020) and its accuracy; the sensitivity of viral RNA swabs has been shown to vary significantly depending on the site, quality and disease stage, ranging from 93% for broncho-alveolar lavage samples to 32% for throat swabs in one study(22). Of those who stated they had not been infected, 84.09% considered themselves at risk, the majority of whom deemed this as high.

41.9% of participants reported suffering from a mental health condition relating to their work during the COVID-19 pandemic. Over a third reported anxiety, a third reported emotional distress, and other disorders including post-traumatic stress disorder and insomnia were also reported. 56.9% experienced anxiety with regards to their colleagues falling ill with COVID-19 and over three quarters of participants (79.63%) denied they wouldavoid going to work during the pandemic. This is arguably a testament to the sense of duty engendered by the participants towards their role as doctors and towards their colleagues. Other studies of healthcare professionals during previous pandemics have found an association between the number and degree of worries with intentional absenteeism, contrary to our study’s findings with this cohort(13). One reason for avoiding workcould include a sense of isolation from social networks. In a previous study during the A/H1N1 influenza pandemic, somehealthcare workers restricted their social contacts and felt isolated by their families and friends due to their work(13). Some may choose to isolate themselves due to fear of transmitting infection, as 94.4% of our participants admitted to socially distancing outside of work. In effect, social distancing may prevent a physical harm but inflict a psychological one. Aside from the potential risk posed to the lives of healthcare professionals, the notion of control may contribute to the psychological burden of working during a pandemic. The concept of control and the loss of this in complex situations may lead clinicians tofeela sense of powerlessness as they feel unable to help. This has been observed in other potentially complex clinical situations(23); it is possible the complexity of a novel infectious disease pandemic may give rise to a similar sense of helplessness, in turn leading to negative emotions and psychologicaldistress.

Formulating solutions according to specific problems may be an effective approach to ensure traineewellbeing and their ability to perform their duties optimally. Listening to the concerns of healthcare professionals and working together to reach local solutions, such as over rest facilities and equipment, may empower them and improve morale.(24)Another important facility to support trainee wellbeing is a psychological support service. While 95.6% of our participants felt psychological support services at work were important during the COVID-19 pandemic, only 62.5% stated they would consider using these at work. This highlights the need to increase awareness of psychological support services outside work, such as the London Deanery Professional Support unit.

The COVID-19 pandemic has affected almost every aspect of life for hospital trainees with medical education being no exception.Almost three quarters of respondents reported a disruption to their local and regional teaching programmes, which was resumed in alternative ways.As many sought to inform themselves independently, through discussions with consultants as well as reading academic papers,the willingness many hospital trainees participating in our study showed towards this endeavour is arguably indicative of their desire to continue their professional development even during the COVID-19 pandemic.

Our study found that the hospital trainee experience during the COVID-19 pandemic was marred by worries primarily for the wellbeing of their loved ones above their own. This was compounded by difficulties socially distancing effectively in the workplace as well as low confidence levels in the efficacy of available PPE, resulting in a large proportion of respondents feeling at risk of being infected with COVID-19. This in turn impacted negatively on their physical and psychological wellbeing.

There were limitations to our study which merit discussion in the interest of future work. We focused on the experience of hospital trainees within one NHS trust, while the experience of trainees in other London NHS trusts and indeed in other heavily impacted parts of the country would enable comparisons of experiences, between hospitals and regions. Indeed, our study explored solely the hospital trainee experience, however future similar studies may include other hospital professional groups to provide a comparison of experiences. As all hospital staff, namely those with direct patient contact, are at risk of infection, ensuring safety for all is a priority in future pandemic responses. Furthermore, while the disproportionate effects of COVID-19 onthose of ethnic minority background has been reported, due to the anonymity we employed in the surveys, our study did not follow through along demographic parameters to enable comparison of the white British and ethnic minority experiences and importantly on rates of infection. Additional avenues to be explored would be reasons for avoiding work as well as for not utilising psychological support services. These in turn would serve to better inform those central to pandemic preparedness planning to support hospital trainees and healthcare workers to perform their roles optimally and to safeguard their wellbeing, thereby contributing to improved clinical outcomes for our patients.

## Conclusion

In conclusion, our study demonstrated that a large proportion of hospital trainees were afflicted by numerous worries while working during the COVID-19 pandemic. Coming from a range of training programmes and walks of life, the physical and psychological health of many were impacted while fulfilling their roles as doctors. Despite this, many reserved their greatest worry for their families and colleagues above themselves; a sense of duty and comradeship appeared to be important motivators.

As the first study on this scale in a major NHS Foundation Trust heavily impacted by the COVID-19 pandemic to investigate the worries and wellbeing of hospital trainees, there remain future avenues to explore. We would be keen to explore reasons for those who would avoid going to work during the COVID-19 pandemic, and the barriers to using psychological support services at work despite almost all respondents agreeing this should be available to all.

As trainees are on the frontline alongsidetheirhospital healthcare professional colleagues, they play a significant role in the fight against the COVID-19 pandemic. The adoption of an open and effective approach towards addressing hospital trainees’ concerns, providing safe working conditions, effective PPE, adequate rest facilities, and psychological support are crucial to ensure their wellbeing, to minimise the costs of sickness and in some cases death, as well as to safeguard the robustness of the NHS response for future pandemics.

## Data Availability

All data from previous studies referred to in the manuscript are referenced accordingly in the manuscript.

**Key messages**

- Physical and psychological health of hospital trainees impacted by working during COVID-19 pandemic
- Greatest worries regarding their families, friends and colleagues over their own wellbeing
- Low levels of confidence of PPE, in use of and ability to practice social distancing
- Engaging hospital trainees in solutions and addressing their concerns crucial fo r ensure trainee wellbeing and morale in future pandemics

## License

The corresponding author has the right to grant on behalf of all authors and does grant on behalf of all authors, an exclusive licence (or non-exclusive for government employees) on a worldwide basis to the BMJ Publishing Group Ltd (“BMJ”), and its Licencees to permit this article (if accepted) to be published in The BMJ’s editions and any other BMJ products and to exploit all subsidiary rights, as set out in our licence.

## Transparency declaration

The lead authors affirm that this manuscript is an honest, accurate, and transparent account of the study being reported; that no important aspects of the study have been omitted; and that any discrepancies from the study as planned (and, if relevant, registered) have been explained.

## Ethics, funding, and sponsorship statements

Ethical approval was not required for this study.

No funding was received for this study.

There were no sponsors for this study.

## Contributorship statement

NA and AK contributed equally to this paper. NA and AK contributed equally to conceptualisation, planning, conduct, analysis, writing, and reporting of the work described in the article.

CS contributed to data analysis and review of the paper.

AB contributed to the resources and administration of the project.

CL provided the overall supervision of the project as well as contributing to the conceptualisation, planning, analysis, and review of the paper.

## Patient and Public Involvement statement

Patients and the public were not involved in any way in our research.

## Other statements

Clinical trial registration was not required for our research.

